# Pathophysiology of SARS-CoV-2: targeting of endothelial cells renders a complex disease with thrombotic microangiopathy and aberrant immune response. The Mount Sinai COVID-19 autopsy experience

**DOI:** 10.1101/2020.05.18.20099960

**Authors:** Clare Bryce, Zachary Grimes, Elisabet Pujadas, Sadhna Ahuja, Mary Beth Beasley, Randy Albrecht, Tahyna Hernandez, Aryeh Stock, Zhen Zhao, Mohamed Al Rasheed, Joyce Chen, Li Li, Diane Wang, Adriana Corben, Kenneth Haines, William Westra, Melissa Umphlett, Ronald E. Gordon, Jason Reidy, Bruce Petersen, Fadi Salem, MariaIsabel Fiel, Siraj M. El Jamal, Nadia M. Tsankova, Jane Houldsworth, Zarmeen Mussa, Wen-Chun Liu, Brandon Veremis, Emilia Sordillo, Melissa R Gitman, Michael Nowak, Rachel Brody, Noam Harpaz, Miriam Merad, Sacha Gnjatic, Ryan Donnelly, Patricia Seigler, Calvin Keys, Jennifer Cameron, Isaiah Moultrie, Kae-Lynn Washington, Jacquelyn Treatman, Robert Sebra, Jeffrey Jhang, Adolfo Firpo, John Lednicky, Alberto Paniz-Mondolfi, Carlos Cordon-Cardo, Mary Fowkes

**Author notes:** These authors contributed equally (Clare Bryce, Zachary Grimes, Elisabet Pujadas).

## Abstract

**BACKGROUND:** Severe Acute Respiratory Syndrome Coronavirus-2 (SARS-CoV-2) and its associated clinical syndrome COVID-19 are causing overwhelming morbidity and mortality around the globe, disproportionately affecting New York City. A comprehensive, integrative autopsy series that advances the mechanistic discussion surrounding this disease process is still lacking.

**METHODS:** Autopsies were performed at the Mount Sinai Hospital on 67 COVID-19 positive patients and data from the clinical records were obtained from the Mount Sinai Data Warehouse. The experimental design included a comprehensive microscopic examination carried out by a team of expert pathologists, along with transmission electron microscopy, immunohistochemistry, RNA in situ hybridization, as well as immunology and serology assays.

**RESULTS:** Laboratory results of our COVID-19 cohort show elevated inflammatory markers, abnormal coagulation values, and elevated cytokines IL-6, IL-8 and TNFα. Autopsies revealed large pulmonary emboli in four cases. We report microthrombi in multiple organ systems including the brain, as well as conspicuous hemophagocytosis and a secondary hemophagocytic lymphohistiocytosis-like syndrome in many of our patients. We provide electron microscopic, immunofluorescent and immunohistochemical evidence of the presence of the virus and the ACE2 receptor in our samples.

**CONCLUSIONS:** We report a comprehensive autopsy series of 67 COVID-19 positive patients revealing that this disease, so far conceptualized as a primarily respiratory viral illness, also causes endothelial dysfunction, a hypercoagulable state, and an imbalance of both the innate and adaptive immune responses. Novel findings reported here include an endothelial phenotype of ACE2 in selected organs, which correlates with clotting abnormalities and thrombotic microangiopathy, addressing the prominent coagulopathy and neuropsychiatric symptoms. Another original observation is that of macrophage activation syndrome, with hemophagocytosis and a hemophagocytic lymphohistiocytosis-like disorder, underlying the microangiopathy and excessive cytokine release. We discuss the involvement of critical regulatory pathways.

## Introduction

Severe Acute Respiratory Syndrome Coronavirus 2 (SARS-CoV-2), the infective agent behind COVID-19, has quickly spread around the globe resulting in substantial morbidity and mortality. As of May 1, 2020, there have been over 3.3 million cases globally including 235,000 deaths. In New York alone, there are over 300,000 patients and nearly 19,000 deaths.^1^

Post-mortem examinations, or autopsies, are the gold standard for the elucidation of the underlying pathophysiology of disease. Despite a rapidly growing body of literature focusing on the clinical impact and molecular microbiology of SARS-CoV-2, autopsy studies are by comparison few and far between. Due to legitimate concerns about the risk of infection and limited personal protective equipment, early postmortem studies have been limited in extent to either gross examination only, post-mortem biopsies, restricted to certain organs or case reports.^2,3,4,5^ A multidisciplinary, integrative post-mortem study documenting the full extent of the disease within a sizeable group is still lacking.

In this comprehensive series, we report the findings from 67 consecutive autopsies of patients who died due to COVID-19 and summarize key histopathological findings within each organ system. We then dive deeper into individual patient stories and integrate our findings with existing literature to advance the discussion of possible disease mechanisms at play.

## Methods

### Laboratory Studies and SARS-CoV-2 testing

Pre-mortem nasopharyngeal swab specimens were placed in universal transport media (UTM) and tested by real-time reverse-transcription-polymerase-chain-reaction (RT-PCR) amplification on the fully automated sample-to-result single well double-target test cobas® 6800 SARS-CoV-2 (Roche Molecular Systems, Branchburg, NJ).

### Autopsy and tissue collection

Consented autopsies were performed at the Mount Sinai Hospital, which performs all autopsies for the Mount Sinai Health System’s seven hospitals in New York City. The autopsies were conducted in a negative pressure room utilizing both PPE and techniques recommended from the current CDC Guidance for Postmortem Specimen Collection.^6^ Extended collection procedures were utilized for autopsies with short post-mortem intervals to support future molecular studies.

### Special Stains and Immunostains

Multiple unstained 3-5 μm-thick sections were cut for special stains and immunohistochemistry (IHC). Special stains were applied according to standard protocols. Antibodies employed for IHC are listed in the Supplementary Methods Table. IHC staining was performed on a Leica Bond III automated stainer except for ACE-II, which was stained on the Discovery Ultra VENTANA systems (Roche).

### RNA in Situ Hybridization

RNA ISH for SARS-CoV-2 performed on a Leica Bond III automated stainer using the RNAscope® SARS-CoV-2 probes for the SARS-CoV-2 S gene encoding the spike protein (catalogue #848561, Advanced Cell Diagnostics, Inc., Hayward, CA) was performed according to the manufacturer’s instructions. Briefly, 4-µm formalin-fixed and paraffin-embedded tissue sections were pretreated with heat and protease prior to hybridization. Tissue sections were hybridized separately with the target probe to detect infected cells. Specific staining signals were identified as brown, punctate dots present in the cytoplasm and/or nucleus. Positive controls were prepared from SARS-CoV-2-infected Vero cell lines (gift from Dr. Florian Krammer, Icahn School of Medicine at Mount Sinai, New York, NY).

### Electron Microscopy

Post-mortem specimens for electron microscopy were placed in 3% buffered glutaraldehyde. Following postfixation in 1% osmium tetroxide, tissues were serially dehydrated and embedded in epoxy resin in standard fashion. One-micron toluidine-stained scout sections were prepared for light microscopic orientation; 80nm ultrathin sections for EM were stained with uranyl acetate and lead citrate and examined in a Hitachi 7650 transmission electron microscope at 80kV.

### Imunofluorescence

Immunofluorescence studies were performed on 4µm thick formalin-fixed paraffin-embedded tissue slides following antigen retrieval in boiling citrate buffer pH6 (Vector labs), blocking for 1 hour at room temperature (10% normal donkey serum (NDS)/0.5% triton-X (TX)), primary antibody incubation overnight at 4°C with anti-SARS-Cov2 nucleocapsid antibody ((Prosci, 35-579, 1:100) and anti-ACE2 antibody (Abcam, ab15348, 1:500) (1% NDS/0.25% TX)), and species-appropriate Cy3- or A488-conjugated secondary antibody incubation for 4 hours at room temperature (1% NDS/0.25% TX). Nuclear counterstain was with DAPI. For secondary only controls, the primary antibody was omitted. Images were visualized on a Zeiss LSM710 confocal microscope. Controls are shown in Supplementary Methods Figure 1.

## Results

### PATIENT CHARACTERISTICS AND LABORATORY DATA

The Mount Sinai Health System (MSHS) has developed a comprehensive, multidisciplinary strategy to test, monitor and better understand SARS-CoV-2 infection and its associated clinical syndrome, COVID-19, in its large and diverse patient population (**Supplementary Figure 1**). As of May 6^th^, 2020, the clinical laboratories had performed 27,251 SARS-CoV-2 tests, yielding 11,106 positive results, antibody serologic testing on 19,471 samples, yielding 10,529 positive results and cytokine profiles on 3,830 samples. MSHS started performing autopsies on COVID-19 positive patients on 3/20/2020, with a total of 67 completed by 4/29/2020. These patients ranged in age from 34 to 94 years (median 69) and exhibited a range of pre-existing conditions and symptom profiles, including hypertension 62.7%, diabetes mellitus 40.3%, coronary artery disease 31.3%, chronic kidney disease 26.7%, asthma 17.9%, heart failure 14.9%, atrial fibrillation 13.4%, obesity 11.9%, co-infections 10.4%, cancer 7.5%, transplantation 7.5%, and COPD 6%. The average length of stay in the hospital was 9.4 days, median of 6 days (information on two patients was not available). A total of 39 patents were intubated, 22 had health care directives not to intubate and the average time to intubation was 5 days. The average time to death from admission was 9.5 days (range of 0 to 61 days) (**Table 1 and Supplementary Figure 2**).

**Table 1:**
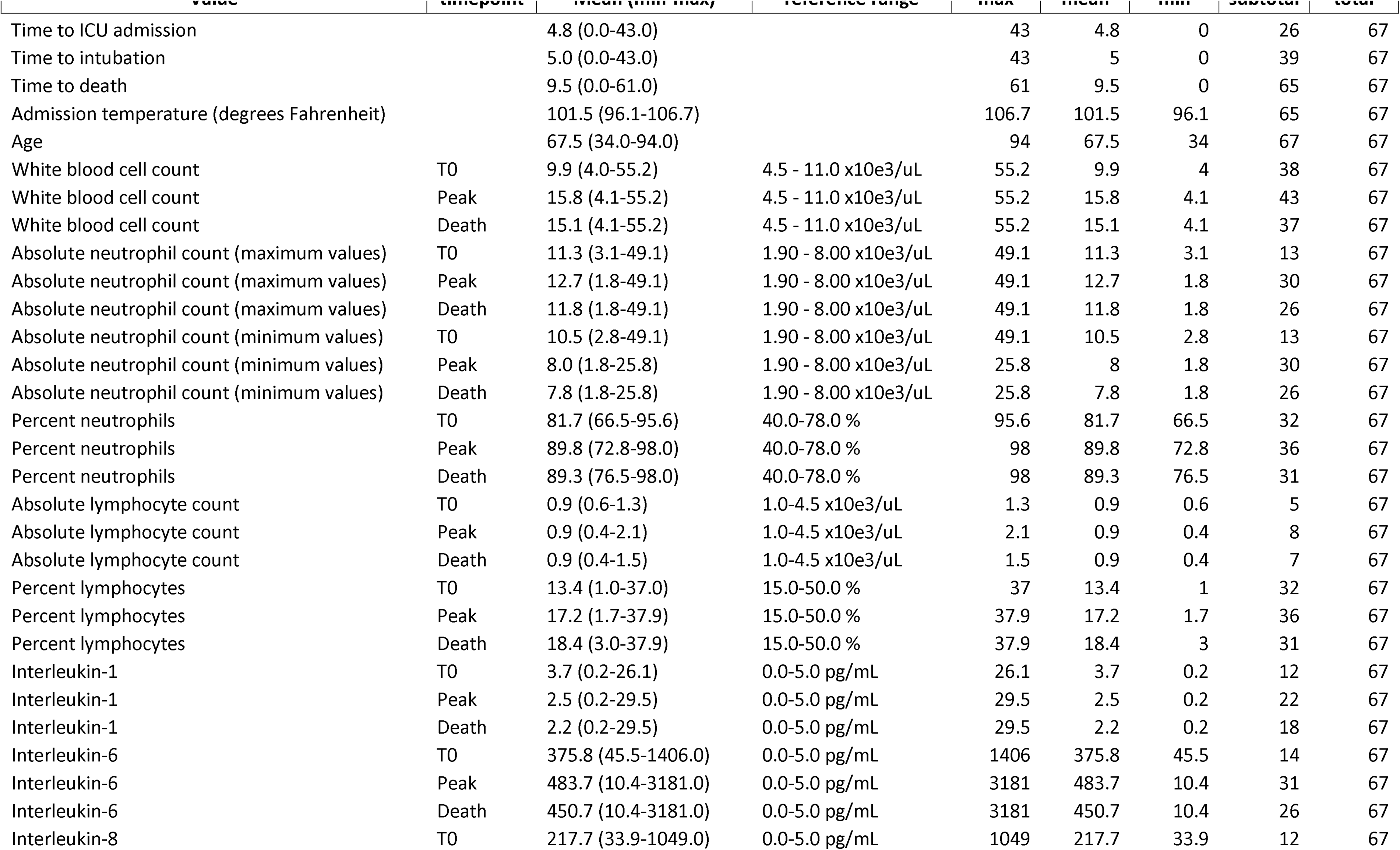
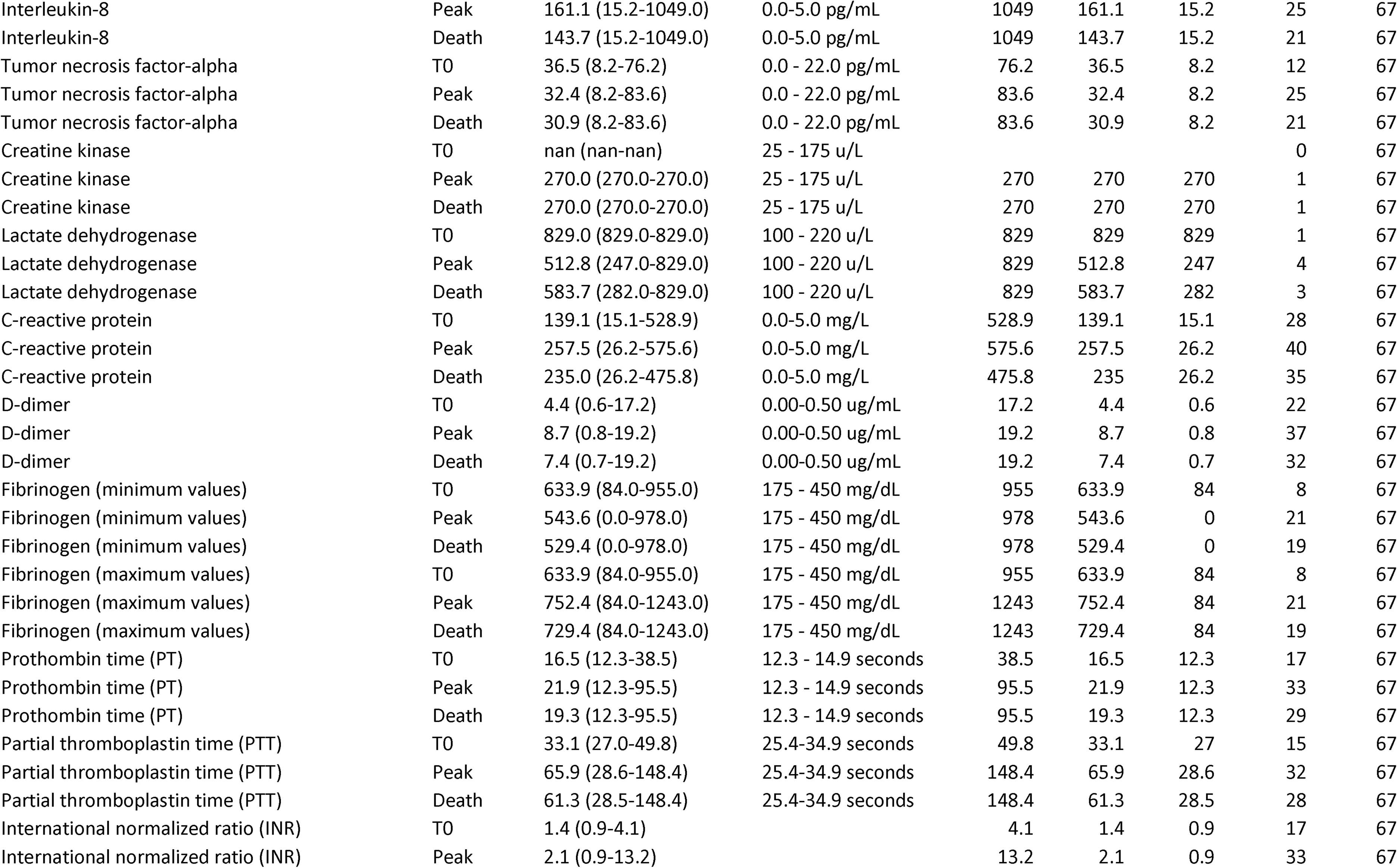
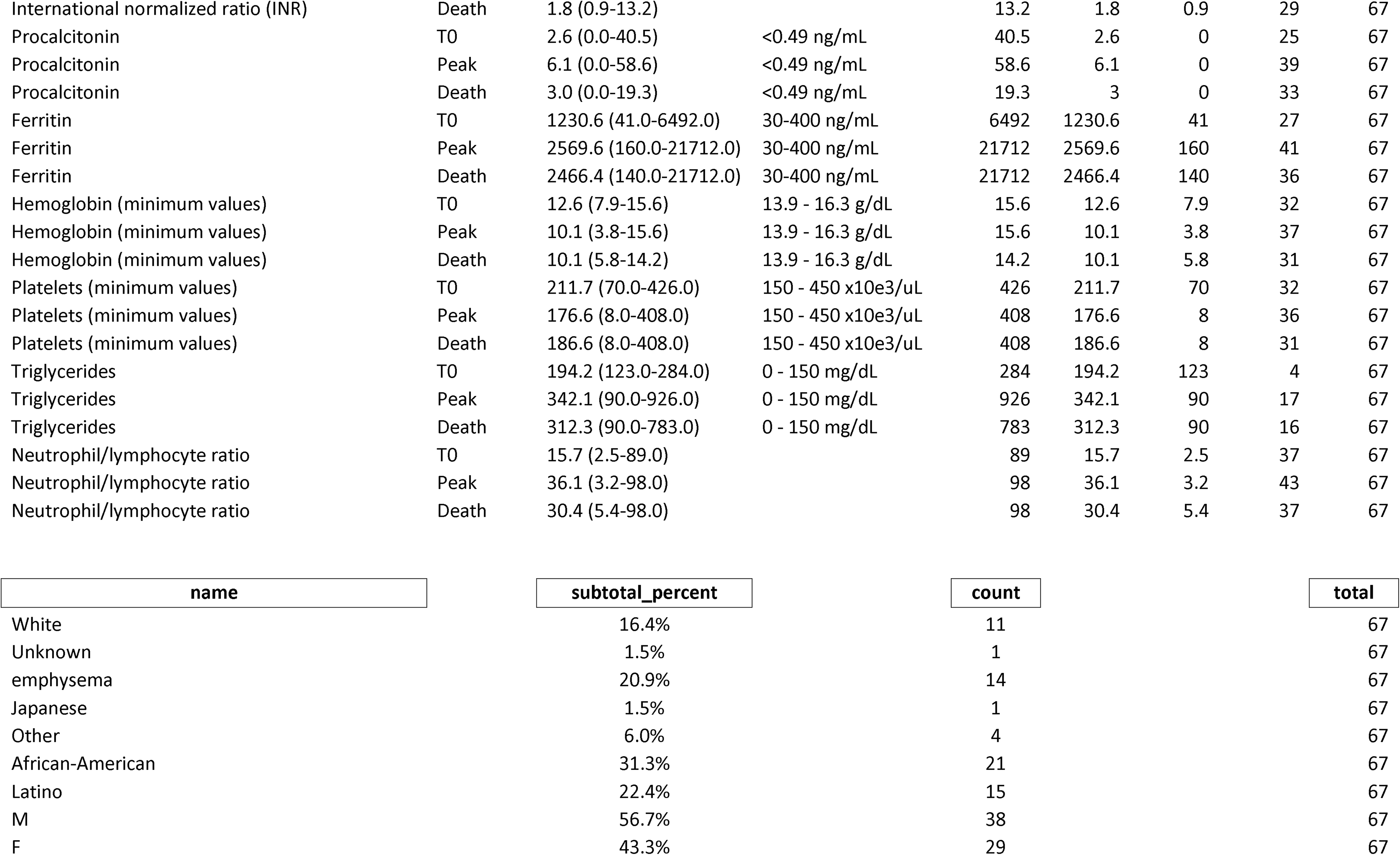
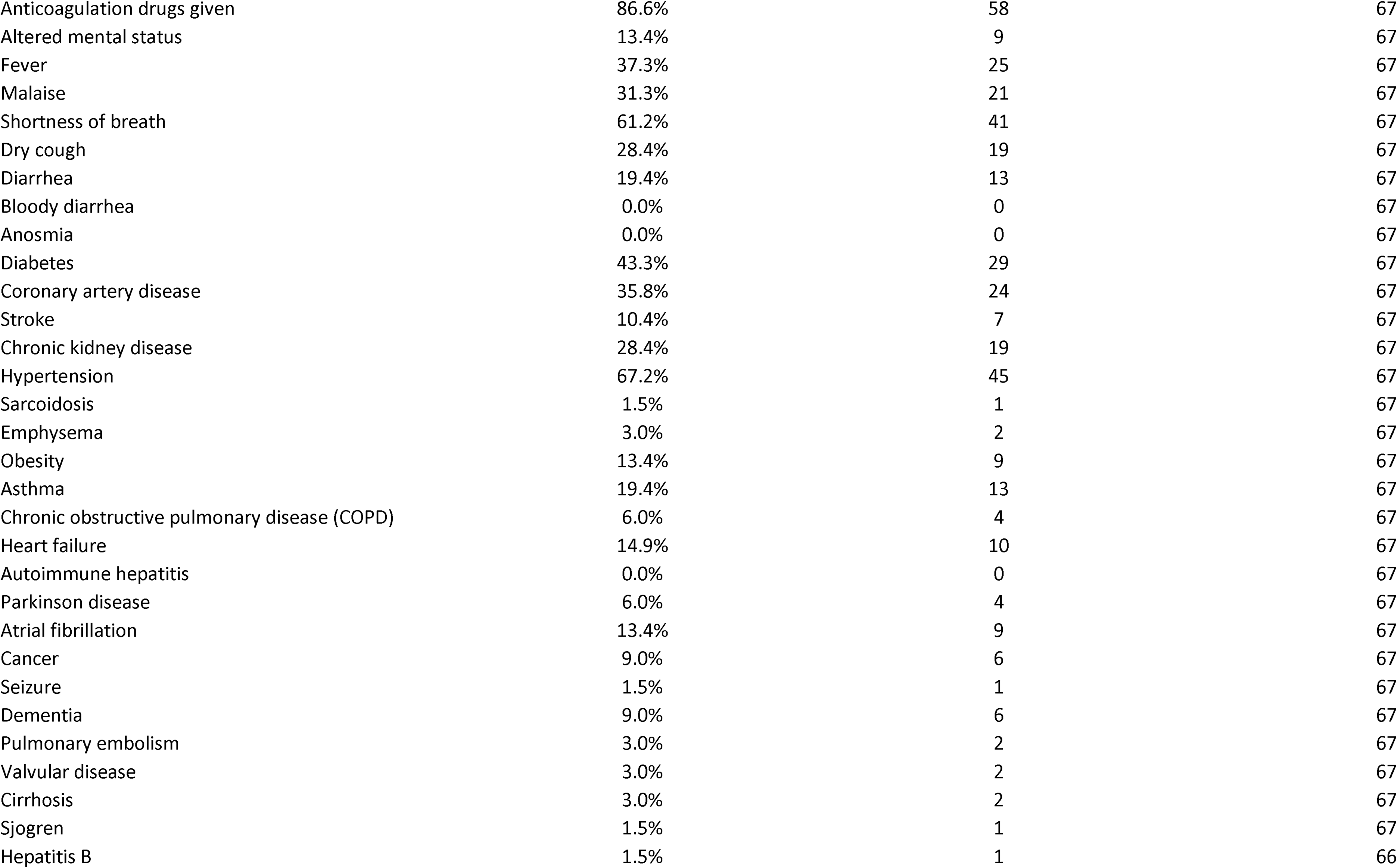
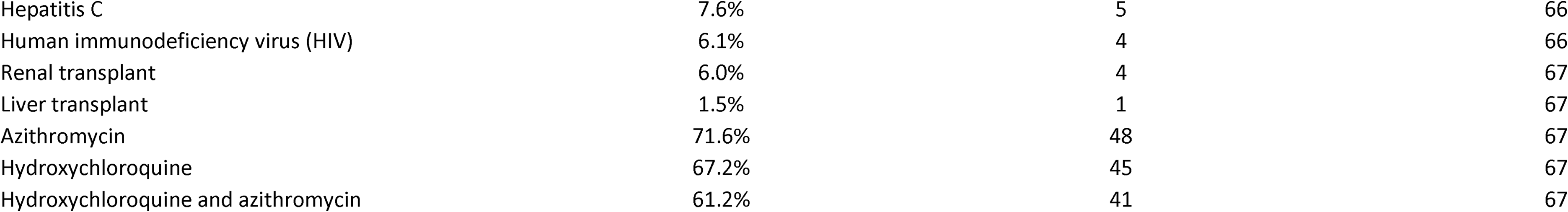
Clinical and Laboratory data summary.

Laboratory values for patients whose autopsies were completed before April 29th were obtained from the Mount Sinai Data Warehouse, which included 49 patients with 34,491 lab values on 613 unique laboratory tests, with values consistent with those previously reported.^7,8,9^ Markers relating to inflammation, including ferritin (mean at peak of 2,570; ref 30-400 ng/ml), C-reactive protein (mean at peak of 257; ref 0.0-5.0 mg/l), procalcitonin (mean at peak of 6.1; ref <0.49 ng/ml), and white blood cell count (mean at peak of 15.8, ref. 1.90-8.00×10^3^/ul) composed predominantly of neutrophils (mean at peak neutrophil to lymphocyte ratio of 36.1, ref. 3.2-98.0), were elevated above normal reference values. Cytokines IL-6 (mean at peak of 483.7; ref 0.0-5.0 pg/ml), IL-8 (mean at peak of 161.1; ref 0.0-5.0 pg/ml), and TNFα (mean at peak of 32.4; ref 0.0-22.0 pg/ml) were also elevated, with cytokine IL-1b remaining within the reference range for most patients (mean at peak 2.5; ref 0.0-5.0 pg/ml).

Alterations in coagulability were also identified resulting in anticoagulation therapy in 80.6% of patients. D-dimer levels were consistently elevated (mean at peak of 8.7 ug/ml) despite administration of anticoagulative therapy. Fibrinogen was also elevated (mean at peak of 752.4; ref 175-450 mg/dl). Thrombocytopenia was uncommon (platelet mean minimum of 176,000; ref 150,000 - 450,000/uL), with values below 100,000 seen in only 6/36 patients. Elevations were seen in prothrombin time (mean at peak of 21.9; ref 12.3-14.9 seconds) partial thromboplastin time (mean at peak of 65.9; ref 25.4-34.9 seconds), and international normalized ratio (mean peak of 2.1). While these could partially be explained by the presence of anticoagulant therapy administered during most of these patients’ admissions, these values were also elevated outside of the normal range at presentation in some patients who were not previously treated with anticoagulants.

### AUTOPSY FINDINGS

All 67 cases were issued a Preliminary Anatomic Diagnosis that integrated gross anatomical findings with the clinical history and laboratory data, with histologic sampling performed on 25, and extended collection for 11 rapid cases and tissue stock saved for future sampling for the remainder. Histologic and molecular examination is ongoing for this cohort, with the number of cases examined for each organ system reported below.

#### Pulmonary findings

The lungs were evaluated in 25 cases, with 10-25 (average 20) slides examined per case. The right lung weight averaged 1013g (range 450-1800g, median 920g) and the left lung weight averaged 886g (range 350-1900g, median 810g) with an appearance ranging from patchy to diffusely consolidated. One case had multiple cavitary lesions. Large pulmonary emboli obstructing the main pulmonary arteries were identified in four cases.

Histologically, the primary finding in the lung parenchyma was diffuse alveolar damage (DAD) in the acute/exudative or early proliferative phase, variably involving 22 cases. Hyaline membranes were present diffusely in 9 cases (**Figure 1A, top left**), with patchy or focal involvement in the remainder. There was associated type 2 pneumocyte hyperplasia with cytologic features of pneumocyte proliferation ranging from bizarre to markedly pleomorphic, generally correlating with the extent of disease. Multinucleated cells were observed in cases with pneumocyte atypia but intranuclear inclusions suggesting viral cytopathic effect were identified in only two cases (**Figure 1A, top right**). Interstitial lymphocytic inflammation was generally mild and diffuse. Seven cases had superimposed acute pneumonia that was extensive and necrotizing in two. Areas of capillary proliferation with inflammation and injury resembling capillaritis was observed in fourteen cases (**Figure 1A, bottom left**). Intravascular fibrin thrombi were observed in seventeen cases within medium sized arteries or arterioles. Platelet aggregates and/or thrombi were difficult to appreciate on H&E stained sections, but CD61 stains performed on 23 of the cases highlighted aggregates or thrombi in medium sized arteries, arterioles and capillaries in all but 2 cases (**Figure 1A, bottom right**). Electron microscopy studies revealed the presence of multiple spherical virus particles showing spike-like electron-dense peplomeric projections ranging from 100-to-140 nm (**Figure 1B**), which was also detected by immunofluorescence studies on the same COVID19-positive patient and further demonstrated focal co-localization of the SARS-CoV-2 antigen (red) in ACE2-positive bronchiolar epithelium (green) (**Figure 1C**). RNA in situ hybridization for the SARS-CoV-2 showed dot-like signals in the lung suggestive of virally infected cells (**Figure 1D**; inset is an infected cell-line control).

**Figure 1:**
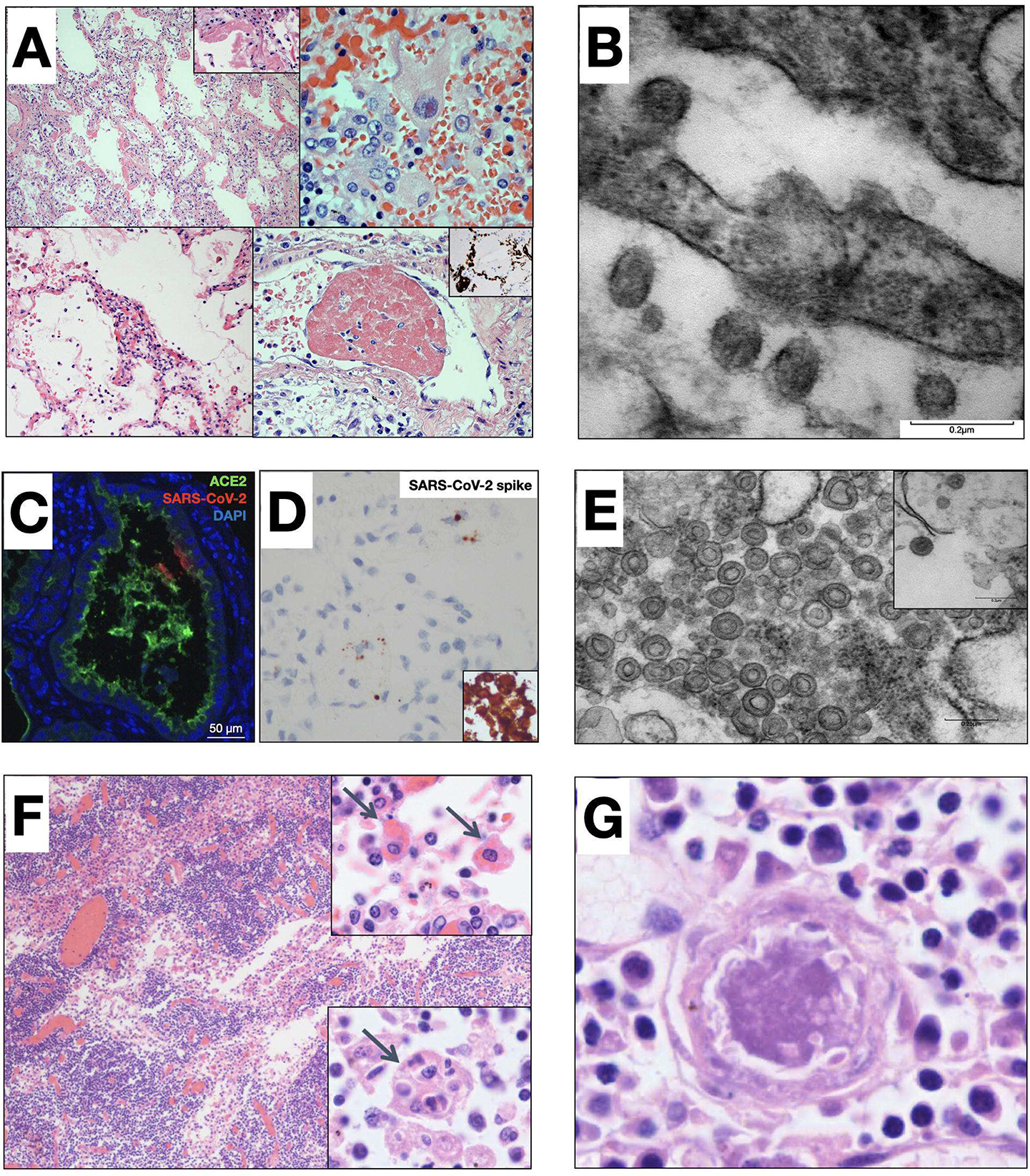
Histologic, IF and EM findings in the lung and thoracic lymph nodes. **(A)** Clockwise from top left. H&E showing diffuse alveolar damage with prominent hyaline membranes (inset, 40x) and pneumocyte hyperplasia (20x); intranuclear inclusions suggestive of viral cytopathic effect (60x); fibrin thrombi in blood vessels (60x), with CD61 staining highlighting platelets (inset, 40x); alveolar septum showing an area of capillary proliferation admixed with chronic inflammatory cells with associated damage (40x). **(B)** Electron micrograph from lung tissue portraying presence of multiple spherical virus particles showing spike-like electron-dense peplomeric projections ranging from 100-to-140 nm. Note the attachment / budding of viral-like particles to the respiratory epithelium signaling possible endo/exocytosis as well as presence of extracellular virus particles. Scale bar, 0.2 μm. **(C)** Immunofluorescence staining in lung tissue showing focal and specific SARS-CoV-2 viral immunoreactivity, including co-localization of viral nucleocapsid antigen in ACE2-positive bronchial epithelium. **(D)** RNAscope SARS-CoV-2 spike gene detected in the lung, with positive controls as insets (single-stranded, top; S-protein, bottom). **(E)** Electron micrograph of a thoracic lymph node revealing coronavirus-induced organelle-like replicative structures consistent with double-membrane vesicles (DMVs), scale bar, 0.25 μm. Inset: Intracytoplasmic spherical shaped virus particles with characteristic electron-dense envelop and fine peplomeric projections, scale bar 0.2 μm. **(F)** Thoracic Lymph nodes containing hemophagocytic histiocytes within sinusoids (10x) with phagocytosed erythrocytes (top insert: 100x) and engulfing multiple cell types (bottom insert: 100x). **(G)** Thoracic Lymph node with microthrombi (H&E, 100x).

#### Lymph node findings

Electron microscopy in the lymph node revealed Coronavirus-induced organelle-like replicative structures consistent with double-membrane vesicles (DMVs) (**Figure 1E**) and intracytoplasmic spherical virus particles with a characteristic electron-dense envelope and fine peplomeric projections. Thoracic lymph nodes showed sinus histiocytosis in 11 cases with focal hemophagocytosis in 9 cases (**Figure 1F**), and multinucleate histiocytes in one. Microthrombi were identified in lymph nodes of 2 cases (**Figure 1G**). Immunohistochemical stains were performed on lymph nodes of three representative cases. One case showed markedly diminished CD3+ T-lymphocytes, with relative preservation of B-cell follicles. The other two cases showed no overt loss of B or T-lymphocytes. All three cases showed predominance of CD4+ T-lymphocytes over CD8+ T-lymphocytes, with CD4+ to CD8+ ratios ranging from approximately 5:1 to 30:1.

#### Cardiovascular findings

The heart was evaluated in 25 cases. All but one had gross cardiac enlargement, with many having left ventricular hypertrophy and moderate to marked atherosclerotic narrowing of the coronary arteries. Histologically, all cases revealed varying degrees of myocyte hypertrophy and interstitial fibrosis, consistent with pre-existing hypertensive and/or atherosclerotic cardiovascular disease. In two cases, there was also patchy mild interstitial chronic inflammation within the myocardium without associated myonecrosis (**Figure 2A**). In 15/25 cases (60%), there was a patchy epicardial mononuclear infiltrate (**Figure 2B, 2C**) that had a predominance of CD4+ T-lymphocytes over CD8+ T-lymphocytes. In three cases, there were also small vessel thrombi in areas where there were epicardial inflammatory infiltrates (**Figure 2B, inset**). In a single case, there was hemophagocytosis within an area of epicardial inflammation (**Figure 2C**), inset.

**Figure 2:**
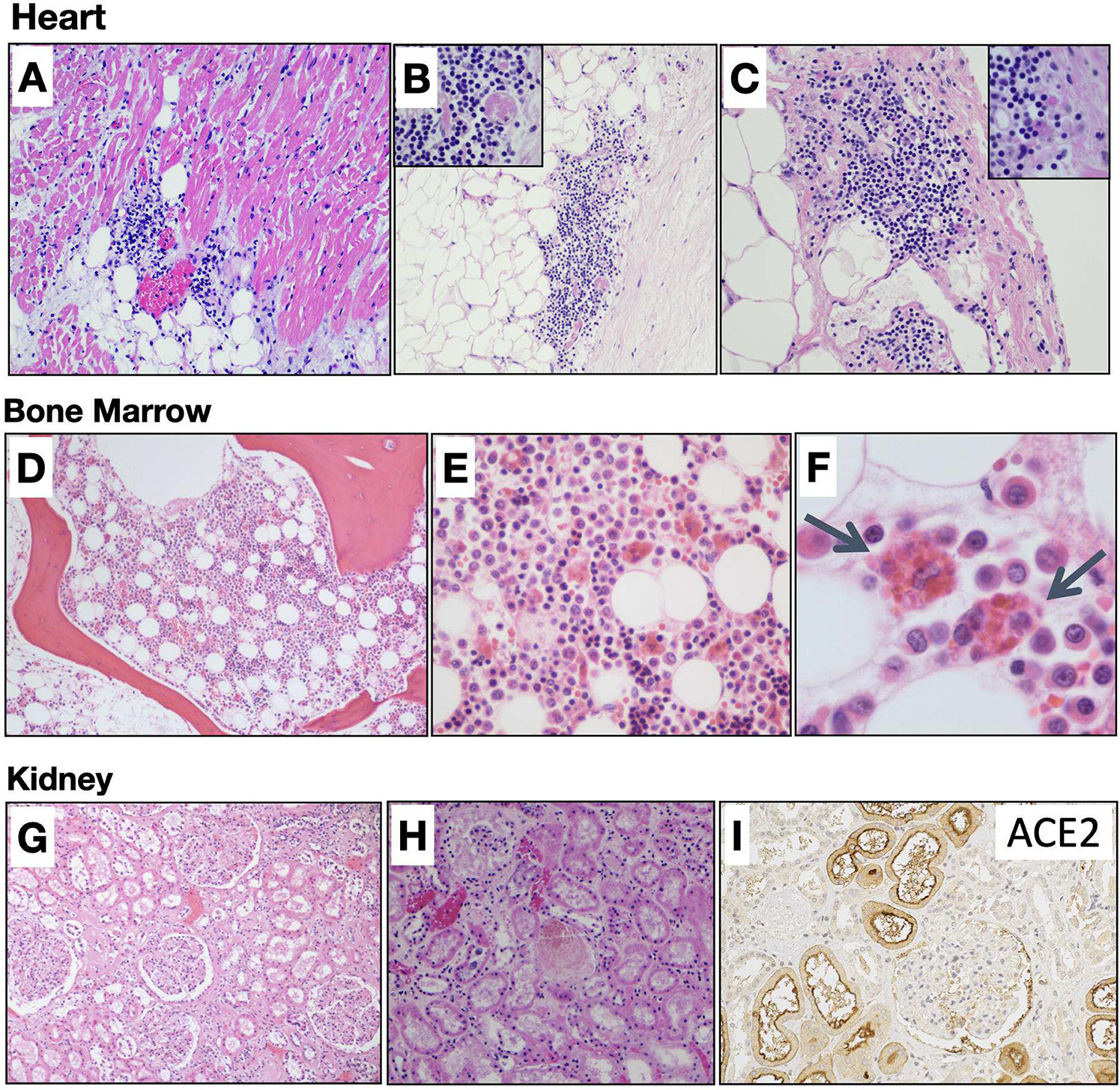
Additional histologic findings. Top Panel. H&E of the heart. **(A)** (10x) Patchy mild interstitial chronic inflammation within the myocardium without myonecrosis. **(B)** (10x) Epicardial mononuclear cell infiltrate with small vessel thrombi (inset at 40x). **(C)** (20x) Epicardial inflammation with focal hemophagocytosis (inset at 40x). Center Panel. H&E of bone marrow at increasing magnifications showing hemophagocytosis. **(D)** 4x. **(E)** 20x **(F)** 100x. Bottom Panel. H&E of the kidney. **(G)** (10x) Glomeruli with mesangial expansion and proximal tubules showing loss of brush borders and attenuation of tubular epithelial cells **(H)** (10x) Tubular injury and intratubular casts in proximal tubules. **(I)** (20x) ACE2 receptors staining was expressed in tubules but not expressed in glomeruli nor endothelial cells.

#### Bone marrow and spleen findings

Hemophagocytosis was also identified in 4/6 bone marrows examined (**Figure 2D-F**, increasing magnification), two of which also showed left-shifted granulopoiesis, one of which showed decreased megakaryocytes, and another one that showed decreased erythroid precursors. The bone marrows were otherwise unremarkable. Spleens from 22 cases were examined. Well-preserved spleens showed hemophagocytic histiocytes, identified in 9 cases. The majority of spleens showed preservation of white pulp, with white pulp nearly absent (<5% of the area of the parenchyma) in two cases. See below for spleen findings in patient 21.

#### Renal findings

The kidney was evaluated in 25 cases, including two with kidney transplants. The most common finding was arterionephrosclerosis. There were four cases that showed glomerular mesangial expansion and two cases that showed mesangial nodular sclerosis, reflecting the prevalence of hypertension and diabetes among this group. Changes suggestive of immune complex deposition in the glomeruli were not seen, though immunofluorescence studies were not performed, nor was collapsing glomerulopathy (Figure 2G). Acute tubular injury was seen diffusely in only two cases. In addition, four cases showed mild patchy tubular injury and fine vacuolization of epithelial cells of proximal tubules (Figure 2H). There was no evidence of viral cytopathic effects found in the tubules. Calcium phosphate crystals were seen in distal tubules of four cases.

One of four patients with postmortem findings of DVT/PE showed thromboemboli. There was frequent congestion of glomerular and peritubular capillaries with focal aggregation of platelets without evidence of thrombotic microangiopathy (TMA). ACE2 receptor staining in the kidney showed expression in apical aspect of proximal tubules and parietal epithelial cells and lack of expression in the glomeruli and endothelial cells (Figure 2I)

#### Liver findings

A total of 22 cases were examined. Macroscopically, two were cirrhotic, two showed nutmeg congestion, one had multiple cysts, and 17 were grossly unremarkable. Ischemic coagulative necrosis involving zone 3 was seen in 5 of 22 (23%) cases with two showing neutrophilic infiltrates similar to ischemia-reperfusion injury. Eight cases showed venous outflow obstruction (VOO), which was acute in five. Scattered within congested areas are Kupffer cells with phagocytosed red blood cells, indicative of hemophagocytosis. Early organizing thrombi staining for CD61 involving portal venules were also seen in 15 cases (68%), which were rarely also found in some hepatic arteries and arterioles.

A significant number of cases had steatosis (15 out of 22, 68%, ranging from mild to severe), indicative of the presence of nonalcoholic fatty liver disease (NAFLD). Masson trichrome stain was performed and revealed seven cases with significant fibrosis. One case with known HCV showed grossly nodular architecture and was confirmed to have stage 3 fibrosis in transition to cirrhosis and two showed established cirrhosis. Five cases showed calcifications in sinusoidal spaces and rarely along the wall of the hepatic arteries. The von Kossa stain was positive in two out of the five cases.

#### Gastrointestinal findings

The gastrointestinal organs were evaluated in 20 cases. Eleven patients had symptoms suggesting gastrointestinal disease including 9 with diarrhea (45%) and 2 with diarrhea and vomiting (10%). The organs were macroscopically unremarkable. Light microscopic examination of 4 specimens of esophagus, 11 of stomach, 16 of small intestine and 17 of colon, excluding 3 autolyzed specimens, revealed no histological abnormalities. None of the patients underwent endoscopic biopsies during their hospitalization.

#### CNS findings

Neuropathological examination of 23 out of 35 brains with histological analysis of 20 revealed a range of abnormal pathology. The most striking findings related to the widespread presence of microthrombi and acute infarction observed in six cases (6/20). The vascular distribution of the infarcts was variable: one case showed a large cerebral artery territory infarct (patient 41) but much more common was small and patchy peripheral and deep parenchymal ischemic infarcts (patient 33, 51, and 46) and others (particularly deep gray matter structures) were hemorrhagic (patient 56). In two of the cases with clinical infarction (patient 41 and 56) there was global anoxic brain injury. In all six cases, a notable and consistent abnormality was the presence of microthrombi often associated with small and patchy infarction. Vascular congestion appeared out of proportion to what is typically seen in routine neuropathological evaluation, and was sometimes accompanied by acute parenchymal microhemorrhages, especially within the necrotic area of infarction, suggestive of vascular damage and reperfusion injury.

The remainder of brains examined, representing the majority of cases, showed less striking histological abnormalities with minimal inflammation and slight neuronal loss without acute hypoxic-ischemic changes. A focal parenchymal infiltrate of T-lymphocytes detected in two cases (patient 14 and 33) was above the normal range, suggesting that focal emerging encephalitis cannot be entirely excluded. Importantly, widespread meningoencephalitis, microglial nodules, and viral inclusions were not a prominent feature in any of the cases, including in the olfactory bulbs and brainstem. There was also no loss of myelin seen on Luxol-Fast-blue/H&E stained sections to suggest demyelination.

### INDIVIDUAL PATIENTS HIGHLIGHT HEMOPHAGOCYTOSIS AND ENDOTHELIAL DYSFUNCTION

#### Patient 21

A female in her 50s with a history of chronic kidney disease, hypertension, and hyperlipidemia was admitted with fatigue, fever, cough, shortness of breath, chest pain, diarrhea and headache and found to be SARS-CoV-2 positive. She developed COVID-19 pneumonia with diffuse alveolar damage and ultimately died of COVID-19 pneumonia with features of congestive heart failure (timeline in **Figure 3A**). Cytokine values were IL6 200-300 (ref. 0-5 pg/mL), IL8 70-80 (ref. 0-5 pg/mL), IL1b 0.5-1.0 (ref 0.5 pg/mL) and TNFa 30-40 (ref 0-22pg/mL). She was found to have cardiomegaly and bilateral pulmonary consolidation. On histology, there were classic findings of DAD (**Figure 3B**), including prominent hyaline membranes. The heart had areas of fibrosis and myocyte hypertrophy consistent with hypertensive changes and prior ischemic insults (**Figure 3C**). The spleen (**Figure 3D**) showed hemophagocytosis (**Figure 3E**) and CD163 positive macrophages (**Figure 3F**). There was a predominance of CD4+ T-lymphocytes over CD8+ T-lymphocytes in the lymph node (Figures 3G and 3H). Renal parenchyma was consistent with known chronic kidney disease (**Figure 3I**). While not fulfilling all the clinical criteria for hemophagocytic lymphohistiocytosis (HLH), this patient had several features in addition to hemophagocytosis on histology, including fever, anemia, and elevated ferritin. Triglycerides were within normal limits.

**Figure 3:**
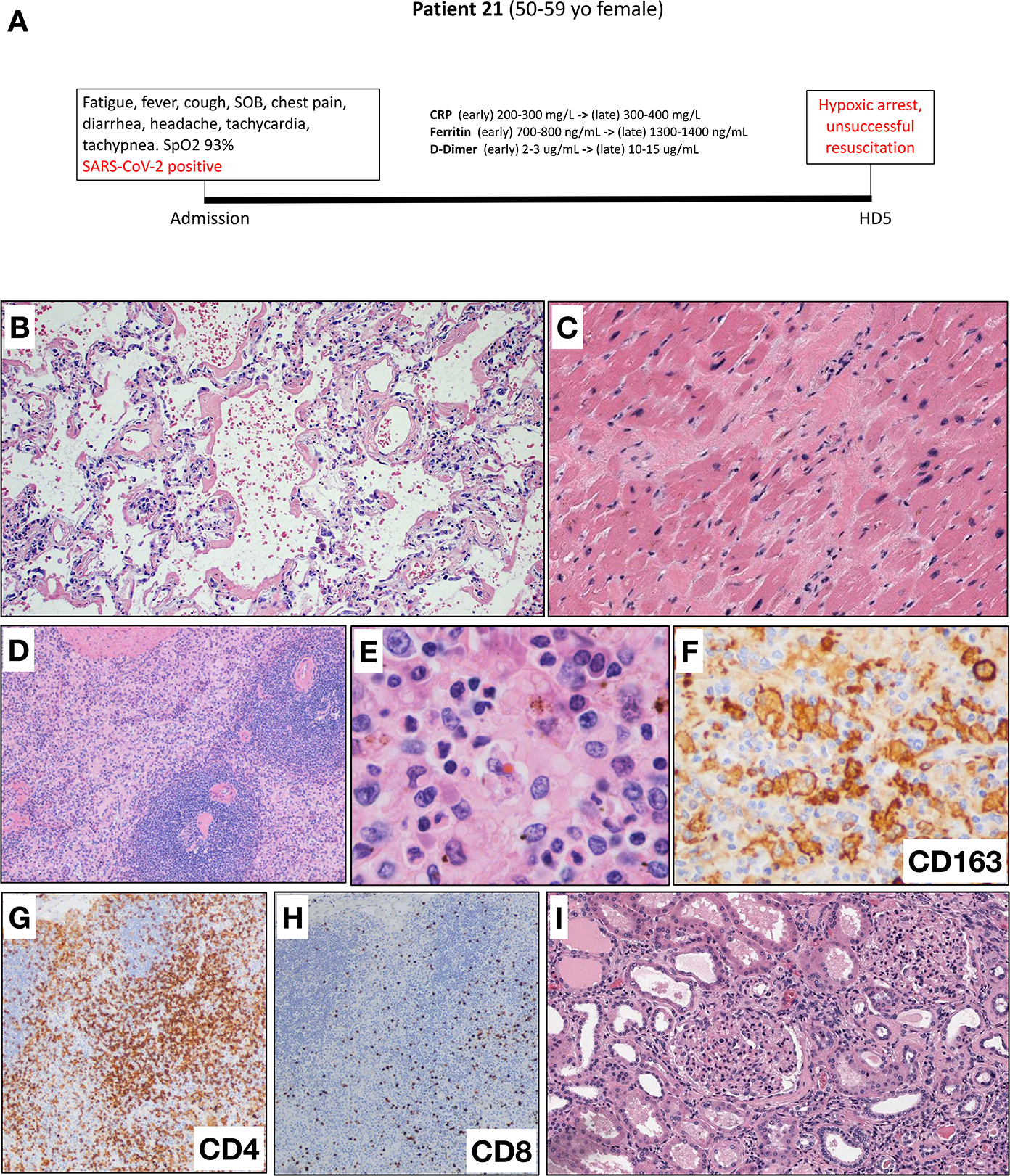
Timeline for patient 21. **(A)** Timeline from admission to death on hospital day 5 with pertinent clinical and laboratory values. **(B)** H&E 4x, lung parenchyma with hyaline membranes. **(C)** H&E 10x, myocardium with fibrosis and myocyte hypertrophy. (**D&E**) H&E 10x and 40x respectively, Spleen with hemophagocytosis. **(F)** CD163 100x, spleen with positive staining of macrophages. **(G&H)** CD4 and CD8 each at 10x respectively in thoracic lymph node, increased CD4 to CD8 positive staining of T-cell lymphocytes. (**I)** H&E 10x, renal parenchyma with known chronic kidney disease.

#### Patient 33

A male in his 60s with a history of hypertension, cancer, an organ transplant, and HIV presented with prolonged fever, chills, a sore throat, shortness of breath, fatigue, night sweats and diarrhea. He had a complicated hospital course (**Figure 4A**) and ultimately died of COVID-19 pneumonia. Cytokine values were not available for this patient. In addition to features of HLH, including fever, anemia, elevated ferritin and splenomegaly, this patient demonstrated multifocal microthrombi with acute infarcts in the frontal, parietal and occipital lobes with associated petechial microhemorrhages. These findings were evident grossly (**Figure 4B**), as well as microscopically, where extensive sampling revealed scattered and sometimes numerous multifocal microthrombi in both leptomeningeal and cortical small-to-medium vessels (**Figure 4C**). Interestingly, the vessels within the brain parenchyma and myocardium stained positive for ACE2, suggesting that they may be a target for SARS-CoV-2 (**Figure 4D**).

**Figure 4:**
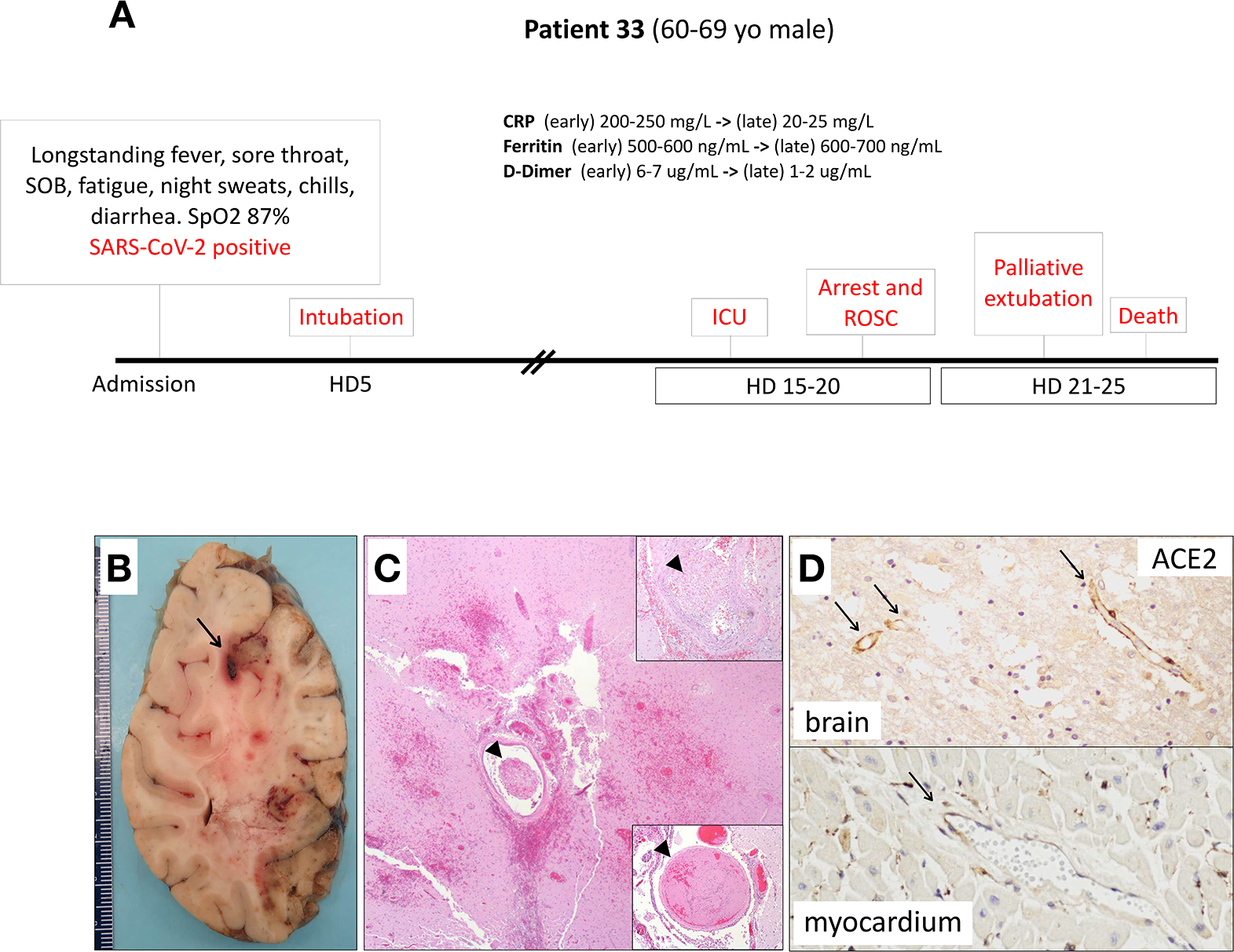
Timeline for patient 33. **(A)** Timeline from admission to death on hospital day 23 with pertinent clinical and laboratory values. **(B)** Coronal section of the right cerebral hemisphere with multiple acute infarcts and focal region suggestive of thrombosed and hemorrhagic blood vessel (arrow). **(C)** Focal region of acute infarct and microhemorrhages with numerous microthrombi (arrowheads). Insets: H&E 10x of two additional identified microthrombi (arrowheads). **(D)** positive ACE2 receptor staining of brain parenchyma blood vessels (10x) and myocardium blood vessels (20x) (arrows).

## Discussion

Conceptualized as a primarily respiratory viral disease that poses a risk to elderly patients with co-morbidities, COVID-19, the clinical syndrome caused by SARS-CoV-2, is increasingly recognized as a far more complex multi-organ and heterogeneous illness than initially anticipated. Some patients display a diverse array of symptoms and complications, including thromboembolic disease even in the setting of anticoagulant therapy, ^10^ a hyperinflammatory state, neuropsychiatric symptoms, and clinical courses complicated by abrupt, unexpected deterioration during the recovery phase. In addition, reports of morbidity in young patients^11^ and children with a Kawasaki-like syndrome^12^ are emerging. Our findings support the notion that endothelial damage and imbalance of both innate and adaptive immune responses, with aberrant macrophage activation, may play a central role in the pathogenesis of the more severe forms of COVID-19.

We report evidence of endothelial dysfunction and a hypercoagulable state, likely through both primary and secondary mechanisms, which elucidates certain clinical symptoms observed in COVID-19 patients, including thromboembolisms and neuropsychiatric features. SARS-CoV-2 is known to gain access to the host cell by binding to the angiotensin-converting enzyme 2 (ACE2) receptor,^13^ which is widely expressed through the human vascular endothelium.^14^ Indeed, recent work suggests that SARS-CoV-2 may invade and even replicate within endothelial cells,^15^ with a recent report showing viral-like particles within the endothelium in the frontal lobe.^16^ Interestingly, our immunohistochemical results reveal distinct ACE2 endothelial phenotypes in specific organs, with ACE2 being highly expressed in lung and brain parenchymal capillaries, but not in the kidney. Interestingly, both lung and brain display multiple thrombotic events, while neither glomeruli nor kidney vessels showed thrombotic microangiopathy. Endothelial cell damage in the brain, leading to neuronal hypoxia, is likely a contributory mechanism to delirium seen in these patients that is out of proportion to their overall clinical course. This would explain such neuropsychiatric symptoms in absence of conspicuous inflammation. Such organ-specific findings are reminiscent of those reported in Hantaviruses of the *Bunyaviridae* family that primarily target the pulmonary and renal vascular beds causing a hemorrhagic fever.^17^ We postulate that SARS-CoV-2 causes endothelial damage by binding ACE2 and misbalancing the renin-angiotensin pathway, dysregulating sphingolipids and activating the ceramide pathway, known to mediate endothelial cell apoptosis in the setting of radiation damage.^18,19,20^ Such injury also generates reactive oxygen species, vasoconstriction and hypoxia, and ultimately the deposition of platelets on an exposed vessel basement membrane initiating the intravascular coagulopathy and multi-organ failure, pathognomonic of severe COVID-19 and death.

The more well-known target of SARS-CoV-2, the epithelial surfaces exhibiting an ACE2 positive phenotype, mainly the respiratory epithelium, revealed extensive viral and inflammatory damage. More specifically, high levels of erythrocyte sedimentation rate (ESR), c-reactive protein (CRP), and cytokines IL-6, IL-8 and TNFα, which are abnormal in our patients, are part of the mechanisms of injury involved in acute respiratory distress syndrome (ARDS) and hypercoagulation^3,21,22^. Similar findings have been previously reported in SARS-CoV-1 and MERS; and have been more recently described in COVID-19 patients.^23,24,25,26^ Among the altered cytokines, significantly elevated IL-6 was a poor prognostic marker in critically ill patients.^27,28^ Furthermore, the endothelium could additionally be damaged via such secondary mechanisms triggered by excessive and uncontrolled release of pro-inflammatory cytokines.

The identification of hemophagocytic histiocytes in lymph nodes (9/11), spleen (9/22), bone marrow (4/6) and other target organs, such as the heart and liver, is consistent with a hyperinflammatory syndrome, and suggestive of secondary hemophagocytic lymphohistiocytosis (sHLH), also known as macrophage activation syndrome. While sHLH is a rare entity, in adults it is most commonly precipitated by viral infection, and is frequently associated with pulmonary complications, including ARDS.^29^ Cytokine abnormalities in COVID-19 patients parallel those seen in sHLH^30^ and suggest a role for the reticuloendothelial system, spleen and bone marrow as important mechanistic sites. Evaluation of lymph nodes showed preservation of B-cells in follicles, while three of four cases showed decreased CD8+ T-lymphocytes, and two showed decreased T-lymphocytes overall. While acknowledging small sample size, these findings are suggestive of histological corollaries to the decrease in peripheral blood lymphocytes seen in our patients, consistent with previous reports suggesting this may be a poor prognostic marker.^31,32^ Our histologic findings of prominent hemophagocytosis and a sHLH-like phenotype in a subset of patients with concomitant elevation in cytokines and inflammatory markers suggest the importance of considering the imbalance of innate and adaptive immune responses with aberrant activation of macrophages, and a blunted adaptive immune response as a core feature in the pathogenesis of severe forms of COVID-19. Extension of these mechanisms to the understanding of the pediatric multi-system inflammatory syndrome potentially associated with COVID-19 is already being pursued.

Overall, our ongoing, systematic and integrative post-mortem study of a large cohort of COVID-19 patients emphasizes the critical role played by endothelial dysfunction and aberrant innate and adaptive immune responses in severe forms of the disease, thus encouraging a shift in therapeutic strategies.

## Data Availability

all data available are included in figures and tables. For any additional information feel free to contact the senior author.

## Conflict of Interest Statement

The authors report no conflicts of interest.

### Ethical/Institutional Review Board Approval

All autopsies were done with written consent from the legal next-of-kin. The Icahn School of Medicine Institution Review Board considers autopsies as non-human subjects (decedents). Approval for this study was provided by the Icahn School of Medicine Grants and Contracts Office.

## Acknowledgments

We thank Valentín Fuster and Adolfo Garcia-Sastre for their guidance and advice. We thank Etty Cortes for her help with tissue banking. We thank Michael Schotsaert, Teresa Lisa Miorin, and A. Aydillo, Irene Ramos-Lopez for direction and help in the collection and processing of microbiological samples. We thank Florian Krammer and Fatima Amanat for sharing formalin-inactivated SARS-CoV-2 infected cell pellets to serve as positive controls. We thank Akm Juber Ahmed, Jin Xu, Alex Fayad, Monika Garcia-Barros, Sara Olson, Rachel Olivares, Tin Htwe Thin and Frances Avila for their support with immunohistochemistry. We thank Ian Hsu for helpful discussion.

## Supplementary Figure Legends

**Supplementary Figure 1:** End-to-end multidisciplinary approach to the treatment of COVID-19 patients in the Mount Sinai Health System

**Supplementary Figure 2:** Full cohort patient timeline

## Supplementary Methods

**Supplementary Methods Table 1:** Antibody Information

**Supplementary Methods Figure 1:** Supplementary Immunofluorescence negative controls. **(A)** Representative SARS-CoV2 and ACE2 co-staining immunofluorescence (IF) image of lung tissue from a COVID-negative patient with pneumonia and similar postmortem time to the COVID-positive patient’s tissue in Figure 1C. **(B-C)** Representative IF images from secondary only controls in the COVID-positive lung tissue shown in Figure 1C.

## Notes

### Competing Interest Statement

The authors have declared no competing interest.

### Funding Statement

This study was supported by the Department of Pathology, Molecular and Cell-Based Medicine at the Icahn School of Medicine at Mount Sinai.

## References

1. Accessed May 1, 2020, at coronavirus.jhu.edu

2. Liu Q, Wang RS, Qu GQ, et al. Gross examination report of a COVID-19 death autopsy. Fa Yi Xue Za Zhi 2020;36:21–3.

3. Xu Z, Shi L, Wang Y, et al. Pathological findings of COVID-19 associated with acute respiratory distress syndrome. Lancet Respir Med 2020;8:420–2.

4. Tian S, Xiong Y, Liu H, et al. Pathological study of the 2019 novel coronavirus disease (COVID-19) through postmortem core biopsies. Mod Pathol 2020.

5. Barton LM, Duval EJ, Stroberg E, Ghosh S, Mukhopadhyay S. COVID-19 Autopsies, Oklahoma, USA. Am J Clin Pathol 2020;153:725–33.

6. Accessed May, 6, 2020, at https://www.cdc.gov/coronavirus/2019-ncov/hcp/guidance-postmortem-specimens.html

7. Zhou F, Yu T, Du R, et al. Clinical course and risk factors for mortality of adult inpatients with COVID-19 in Wuhan, China: a retrospective cohort study. Lancet 2020;395:1054–62.

8. Zhang Y, Xiao M, Zhang S, et al. Coagulopathy and Antiphospholipid Antibodies in Patients with Covid-19. N Engl J Med 2020;382:e38.

9. Connors JM, Levy JH. COVID-19 and its implications for thrombosis and anticoagulation. Blood 2020.

10. Klok FA, Kruip M, van der Meer NJM, et al. Incidence of thrombotic complications in critically ill ICU patients with COVID-19. Thromb Res 2020.

11. Oxley TJ, Mocco J, Majidi S, et al. Large-Vessel Stroke as a Presenting Feature of Covid-19 in the Young. N Engl J Med 2020.

12. Jones VG, Mills M, Suarez D, et al. COVID-19 and Kawasaki Disease: Novel Virus and Novel Case. Hosp Pediatr 2020.

13. Hoffmann M, Kleine-Weber H, Schroeder S, et al. SARS-CoV-2 Cell Entry Depends on ACE2 and TMPRSS2 and Is Blocked by a Clinically Proven Protease Inhibitor. Cell 2020;181:271–80 e8.

14. Hamming I, Timens W, Bulthuis ML, Lely AT, Navis G, van Goor H. Tissue distribution of ACE2 protein, the functional receptor for SARS coronavirus. A first step in understanding SARS pathogenesis. J Pathol 2004;203:631–7.

15. Varga Z, Flammer AJ, Steiger P, et al. Endothelial cell infection and endotheliitis in COVID-19. Lancet 2020;395:1417–8.

16. Paniz-Mondolfi A, Bryce C, Grimes Z, et al. Central Nervous System Involvement by Severe Acute Respiratory Syndrome Coronavirus -2 (SARS-CoV-2). J Med Virol 2020.

17. Peters CJ, Simpson GL, Levy H. Spectrum of hantavirus infection: hemorrhagic fever with renal syndrome and hantavirus pulmonary syndrome. Annu Rev Med 1999;50:531–45.

18. Ghidoni R, Caretti A, Signorelli P. Role of Sphingolipids in the Pathobiology of Lung Inflammation. Mediators Inflamm 2015;2015:487508.

19. Matsunaga F. [In memoriam: Prof. Toshio Kurokawa]. Nihon Shokakibyo Gakkai Zasshi 1988;85:1217–22.

20. Garcia-Barros M, Paris F, Cordon-Cardo C, et al. Tumor response to radiotherapy regulated by endothelial cell apoptosis. Science 2003;300:1155–9.

21. Huppert LA, Matthay MA, Ware LB. Pathogenesis of Acute Respiratory Distress Syndrome. Semin Respir Crit Care Med 2019;40:31–9.

22. Zhang W, Zhao Y, Zhang F, et al. The use of anti-inflammatory drugs in the treatment of people with severe coronavirus disease 2019 (COVID-19): The Perspectives of clinical immunologists from China. Clin Immunol 2020;214:108393.

23. Hwang DM, Chamberlain DW, Poutanen SM, Low DE, Asa SL, Butany J. Pulmonary pathology of severe acute respiratory syndrome in Toronto. Mod Pathol 2005;18:1–10.

24. Huang C, Wang Y, Li X, et al. Clinical features of patients infected with 2019 novel coronavirus in Wuhan, China. Lancet 2020;395:497–506.

25. Teijaro JR, Walsh KB, Cahalan S, et al. Endothelial cells are central orchestrators of cytokine amplification during influenza virus infection. Cell 2011;146:980–91.

26. Pedersen SF, Ho YC. SARS-CoV-2: a storm is raging. J Clin Invest 2020;130:2202–5.

27. Wu C, Chen X, Cai Y, et al. Risk Factors Associated With Acute Respiratory Distress Syndrome and Death in Patients With Coronavirus Disease 2019 Pneumonia in Wuhan, China. JAMA Intern Med 2020.

28. Ruan Q, Yang K, Wang W, Jiang L, Song J. Clinical predictors of mortality due to COVID-19 based on an analysis of data of 150 patients from Wuhan, China. Intensive Care Med 2020.

29. Mehta P, McAuley DF, Brown M, et al. COVID-19: consider cytokine storm syndromes and immunosuppression. Lancet 2020;395:1033–4.

30. McGonagle D, Sharif K, O’Regan A, Bridgewood C. The Role of Cytokines including Interleukin-6 in COVID-19 induced Pneumonia and Macrophage Activation Syndrome-Like Disease. Autoimmun Rev 2020: 102537.

31. Wang F, Nie J, Wang H, et al. Characteristics of peripheral lymphocyte subset alteration in COVID-19 pneumonia. J Infect Dis 2020.

32. Chen N, Zhou M, Dong X, et al. Epidemiological and clinical characteristics of 99 cases of 2019 novel coronavirus pneumonia in Wuhan, China: a descriptive study. Lancet 2020;395:507–13.

